# Reliability of retinal pathology quantification in age-related macular degeneration: Implications for clinical trials and machine learning applications

**DOI:** 10.1101/2020.10.09.20210120

**Authors:** Philipp L. Müller, Bart Liefers, Tim Treis, Filipa Gomes Rodrigues, Abraham Olvera-Barrios, Bobby Paul, Narendra Dhingra, Andrew Lotery, Clare Bailey, Paul Taylor, Clarisa I. Sánchez, Adnan Tufail

## Abstract

**Purpose:** To investigate the inter-reader agreement for grading of retinal alterations in age-related macular degeneration (AMD) using a reading center setting.

**Methods:** In this cross-sectional case series, spectral domain optical coherence tomography (OCT, Topcon 3D OCT, Tokyo, Japan) scans of 112 eyes of 112 patients with neovascular AMD (56 treatment-naive, 56 after three anti-vascular endothelial growth factor injections) were analyzed by four independent readers. Imaging features specific for AMD were annotated using a novel custom-built annotation platform. Dice score, Bland-Altman plots, coefficients of repeatability (CR), coefficients of variation (CV), and intraclass correlation coefficients (ICC) were assessed.

**Results:** Loss of ellipsoid zone, pigment epithelium detachment, subretinal fluid, and Drusen were the most abundant features in our cohort. The features subretinal fluid, intraretinal fluid, hypertransmission, descent of the outer plexiform layer, and pigment epithelium detachment showed highest inter-reader agreement, while detection and measures of loss of ellipsoid zone and retinal pigment epithelium were more variable. The agreement on the size and location of the respective annotation was more consistent throughout all features.

**Conclusions:** The inter-reader agreement depended on the respective OCT-based feature. A selection of reliable features might provide suitable surrogate markers for disease progression and possible treatment effects focusing on different disease stages.

**Translational Relevance:** This might give opportunities to a more time- and cost-effective patient assessment and improved decision-making as well as have implications for clinical trials and training machine learning algorithms.

## INTRODUCTION

Age-related macular degeneration (AMD) is a leading cause of legal blindness in the industrialized world.^1^ Concerning advanced disease manifestations, a dry stage defined by the presence of retinal pigment epithelial (RPE) and outer retinal atrophy (RORA, also called ‘geographic atrophy’, GA) can be distinguished from or complicated by a neovascular (nAMD) form typically characterized by the presence of choroidal neovascularization (CNV).^2–4^

While both forms of late stage AMD are associated with the risk of visual loss, an effective treatment for RORA development and progression is still pending. However, various therapeutic approaches are tested in different stages of preclinical and clinical trials.^5,6^ In order to accelerate clinical testing, meaningful, validated clinical endpoints are needed.^7^ Most interventional trials currently rely on the progression of RORA which is an accepted endpoint by regulators.^8,9^ However, the most effective upcoming therapeutic approach might be directed to earlier disease stages.^10^ Therefore, ideal surrogate markers should identify early disease-associated alterations before the hitherto unknown point-of-no return.^11^ A current paper dealing with RORA in mitochondriopathies described a consistent sequence of optical coherence tomography (OCT)-based imaging features in the development of RORA representing different disease stages.^12^ As an international consensus, the Classification of Atrophy Meetings (CAM) group not only defined complete RORA but also reported preceding OCT-features for AMD.^13,14^ However, the reliability of the detection and quantification of some of these features has not yet been systematically and comprehensively investigated. Nevertheless, they have already been implemented by reading centers for current and upcoming observational and interventional trials.^13,15,16^

Concerning nAMD, the therapy with intraocular injection of anti-vascular endothelial growth factor (anti-VEGF) has been shown to be effective and reduces the risk of visual loss.^17,18^ However, the numbers and costs of required visits mean a significant burden on health-care systems, medical personal, and patients, in particular in the light of growing numbers due to demographic changes and rising life expectation.^19^ Therefore, personalized interval and treatment strategies (i.a. ‘Treat & Extend’) are used more commonly in current clinical settings.^20,21^ In this context, objective and reliable features to determine disease activity are crucial. OCT is typically used for monitoring as it provides cross-sectional images of the retina that allow to identify presence as well as extent of these features.^22,23^ Usually, the feature identification is manually performed by human investigators. Machine learning applications are progressively entering this field, especially in the context of potential deployment of in home or remote OCT monitoring.^24^ However, the “gold-standard” by which these algorithms are trained and validated is conventionally human grading. This might raise the question concerning reliability, subjectivity, and bias of the treatment decisions.^25^

In this study, we therefore investigate the reliability of the grading of defined OCT features commonly found in the development of RORA and/or in the presence of CNV secondary to AMD in order to provide estimates for human inter-reader agreement for each of these features. Thereby, we focus on the detection, as well as the size and the overlap of the particular annotations.

## METHODS

This retrospective cross-sectional case series was performed at the Moorfields Eye Hospital NHS Foundation Trust, London, UK. To identify AMD patients, we linked the diagnosis to OCT images of the electronic medical records database (Medisoft, Leeds, UK) of five centers in the United Kingdom using pseudonymized identifiers. The imaging data comprised 6 x 6 mm foveal centered OCT volume scans obtained by spectral-domain OCT (Topcon, Tokyo, Japan). Any other additional ocular pathology (including prior clinically significant macular oedema), prior unlicensed Bevacizumab injections, intraocular surgery within 90 days, or prior macular or panretinal photocoagulation led to exclusion. Thereby, this study included imaging data of 112 eyes of 112 AMD patients at different disease stages. Half of these eyes were treatment-naive, the others were imaged after three anti-vascular endothelial growth factor (VEGF) injections. The study was in adherence with the declaration of Helsinki. The Institutional Review Board ruled that approval was not required for this study, because all data was completely anonymized before being released to research.

### Image analysis

In order to assess the reliability of grading retinal alterations in AMD, a single OCT B-scan per eye was randomly selected for annotation. The other B-scans were available to give additional context if needed. Annotations were performed by four independent trained retinal specialists masked to the results of each other using a custom-build platform (Supplementary Figure S1). All retinal abnormalities were to be delineated. The platform provided default labels for the most common abnormalities (including those described by the CAM group)^13,14^ and allowed the readers to add additional labels not covered by the default setup. Depending on the feature, it was annotated either as area, lateral extent, or number (i.e., single dots in features with pointwise presentation), likewise for all readers. Preset default labels included drusen, loss of ellipsoid loss (EZ), hyperreflective dots (HRD), hypertransmission of OCT-signal (HT), hyporeflective wedges, intraretinal fluid (IRF), descent of outer plexiform layer (OPL), outer retinal tubulations, pigment epithelial detachment (PED), loss of retinal pigment epithelium (RPE), reticular pseudodrusen (RPD), subretinal fluid (SRF), subretinal hyperreflective material (SRHM), and sub-RPE plaques (Supplementary Figure S1).

The annotated images were then evaluated using Python (version 3.8.2). In order to obtain the area measures in mm^2^ and lateral extent measures in mm, the extracted values of annotated features (i.e., in pixels^2^ and pixels) were multiplied by the individual scaling factor depending on the scanning protocol. Further statistical analysis was exclusively made for features present in at least 20 annotated B-scans respectively eyes to ensure reliable results.

### Statistical analysis

The software environment R (version 4.0.2, The R Foundation for Statistical Computing, Vienna, Austria)^26^ was used for inter-reader correlations. To compare the reliability of feature detection, Fleiss coefficients were used.^27^ To measure the agreement in the annotated feature size, lateral extent or number, intraclass correlation coefficients (ICC, one-way random), 95% coefficients of repeatability (CR) and coefficients of variation (CV) were determined.^28–30^ To account for the unbalanced number of readings per sample, a linear mixed-effects model was used. Bland-Altman plots were generated from slices with annotations of at least two readers for visualization of limits of agreement. Spearman’s rank correlation coefficients (ρ) were calculated between the absolute differences and the mean values to evaluate whether measurement variability increases with lesion size or number.^29^

To measure overlap in annotated areas, we calculated the Dice similarity metric using Python (version 3.8.2) whenever more than one reader annotated the same feature within a respective B-scan. It is defined as the size of the intersection of two areas divided by their average individual size, ranging from 0 (indicating no spatial overlap) to 1 (indicating complete overlap).^31^ For area measures, overlap was calculated on voxel-level. For lateral extent measures, only the lateral location of the feature was taken into account. The mean Dice-coefficients per feature is reported. Due to their focal nature, the Dice-coefficient was not regarded an appropriate metric for annotations of HRD.

## RESULTS

In 111 out of the included 112 OCT B-scans, at least one pathologic feature was annotated. Hyporeflective wedges, outer retinal tubulations, RPD and sub-RPE plaques were present but excluded from analysis due to their rarity in the respective scans (presence in less than 20 annotated B-scans). In total, ten features were used for further analysis (Table 1). Out of the latter group, EZ-loss, Drusen, and PED were the most abundant features.

**Table 1:** Inter-reader agreement of feature detection.

| **Grading parameter** | **n** | **Kappa-Coefficient** | **95% CI** |
| --- | --- | --- | --- |
| Drusen | 85 | 0.367 | 0.292 – 0.443 |
| EZ-loss | 108 | 0.260 | 0.185 – 0.336 |
| HRD | 71 | 0.422 | 0.246 – 0.497 |
| HT | 29 | 0.746 | 0.671 – 0.822 |
| IRF | 50 | 0.621 | 0.545 – 0.696 |
| OPL-descent | 20 | 0.611 | 0.536 – 0.687 |
| PED | 77 | 0.598 | 0.522 – 0.674 |
| RPE-loss | 76 | 0.160 | 0.085 – 0.236 |
| SRF | 45 | 0.823 | 0.747 – 0.898 |
| SRHM | 51 | 0.357 | 0.282 – 0.433 |

CI = Confidence interval, EZ = Ellipsoid loss, HRD = Hyperreflective dots, HT = Hypertransmission of OCT-signal, IRF = Intraretinal fluid, n = overall number of B-scans annotated with the respective feature by at least one reader, OPL = Outer plexiform layer, PED = Pigment epithelial detachment, RPE = Retinal pigment epithelium, SRF = Subretinal fluid, SRHM = subretinal hyperreflective material

The feature detection at the B-scan level (i.e., the individual lesion level is important when investigating progression) revealed variable inter-reader agreement (Table 1). The most reliable results could be found in SRF and IRF, that account to neovascular complications, as well as the features HT, OPL-descent, and PED. Only slight to moderate inter-reader agreement could be found in the detection of EZ-loss and RPE-loss.^27^

The evaluation of inter-reader agreement concerning the size, lateral extension or number of annotated features at the B-scan level revealed more consistent results. All ICC values ranged from moderate to excellent correlation (Table 2).^32^ The focality (i.e., number of individual annotated spots) measures of HRD revealed the lowest ICC with values over 0.50. The features with the highest scores for inter-reader agreement of annotated size, lateral extension or number were PED, SRF, HT and OPL-descent in our cohort (ICC > 0.85, Figure 1).

**Table 2:** Inter-reader agreement of size, lateral extension or number of annotated features.

| **Grading parameter** | **CoR** | **CV [%]** | **ICC (95% CI)** |
| --- | --- | --- | --- |
| Drusen | 0.098 * | 55.0 | 0.687 (0.534 – 0.792) |
| EZ-loss | 3.446 † | 42.4 | 0.573 (0.415 – 0.695) |
| HRD_Focality | 9.388 | 64.5 | 0.527 (0.267 – 0.699) |
| HT | 0.625 † | 24.1 | 0.936 (0.880 – 0.968) |
| IRF | 0.121 * | 81.8 | 0.713 (0.525 – 0.831) |
| OPL-descent | 0.763 † | 16.2 | 0.884 (0.739 – 0.952) |
| PED | 0.134 * | 17.6 | 0.972 (0.959 – 0.981) |
| RPE-loss | 2.157 † | 44.8 | 0.614 (0.345 – 0.766) |
| SRF | 0.103 * | 46.5 | 0.938 (0.900 – 0.964) |
| SRHM | 0.234 * | 53.9 | 0.793 (0.644 – 0.880) |

\* = values indicate mm<sup>2</sup>, † = values indicate mm
CI = Confidence interval, CoR = 95% Coefficients of repeatability, CV = Coefficients of variation, EZ = Ellipsoid loss, HRD = Hyperreflective dots, HT = Hypertransmission of OCT-signal, ICC = Intraclass correlation coefficients, IRF = Intraretinal fluid, OPL = Outer plexiform layer, PED = Pigment epithelial detachment, RPE = Retinal pigment epithelium, SRF = Subretinal fluid, SRHM = subretinal hyperreflective material

**Figure 1:**
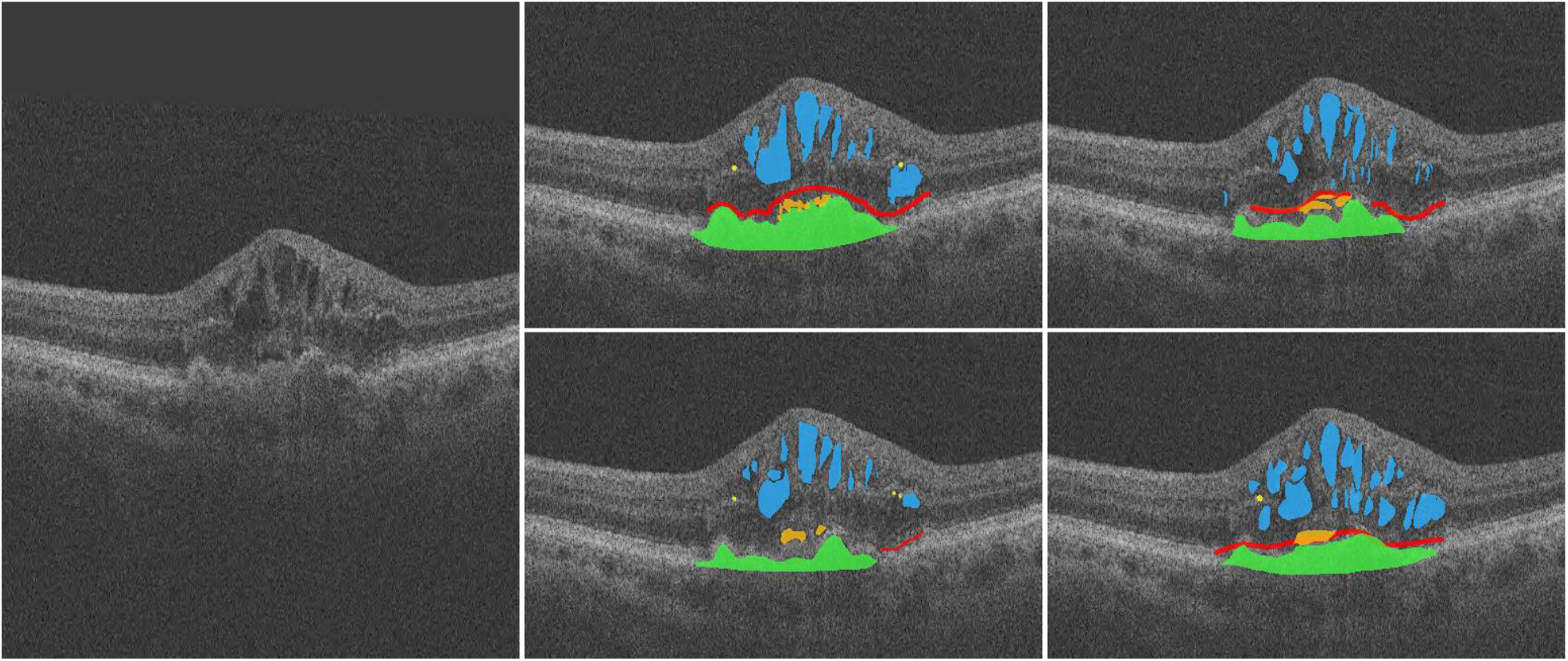
Optical coherence tomography (OCT)-based feature annotation. An OCT B-scan (left) and the respective feature annotation of each reader (right) is demonstrated as example. Intraretinal fluid (IRF, blue), subretinal fluid (SRF, orange), and pigment epithelial detachment (PED, green) revealed high inter-reader agreement, while annotations of loss of ellipsoid zone (EZ-loss, red) and hyperreflective dots (HRD, yellow) significantly differed in size and number between the readers. However, the location of annotated features within the B-scan was quite similar throughout all features.

The Bland-Altman plots did not reveal systematic inter-reader discrepancies (Figure 2 and Supplementary Figures S2 to S11). However, the inter-reader variability increased with annotated area or number according to Spearman’s rank correlation coefficient (ρ) for absolute differences and mean values for measures of Drusen (ρ = 0.317 to ρ = 0.828, P < 0.001 to P = 0.049), PED (ρ = 0.316 to ρ = 0.605, P < 0.001 to P = 0.042), and HRD (ρ = 0.509 to ρ = 0.761, P < 0.001 to P = 0.018). The area measures of IRF (ρ = 0.311 to ρ = 0.755, P < 0.001 to P = 0.139), SRF (ρ = 0.326 to ρ = 0.517, P = 0.003 to P = 0.062), and SRHM (ρ = 0.150 to ρ = 0.436, P = 0.170 to P = 0.708), as well as lateral distance measures of EZ-loss (ρ = 0.010 to ρ = 0.297, P = 0.021 to P = 0.936), HT (ρ = 0.021 to ρ = 0.550, P = 0.027 to P = 0.921), OPL-descent (ρ = 0.036 to ρ = 0.455, P = 0.066 to P = 0.964), and RPE-loss (ρ = 0.108 to ρ = 0.748, P < 0.001 to P = 0.818) did not show this correlation.

**Figure 2:**
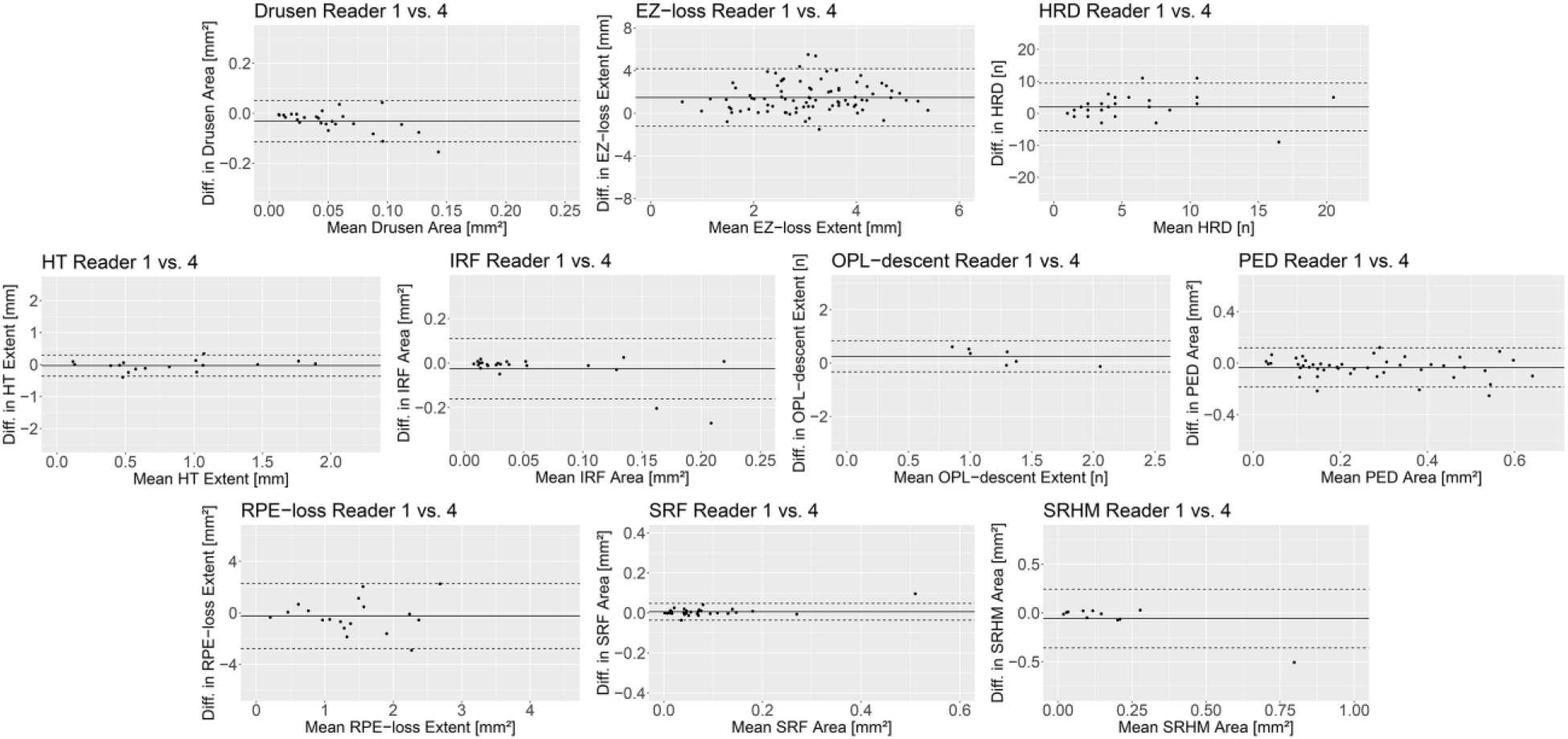
Inter-reader agreement. The Bland-Altman plots demonstrate the inter-reader agreement between two exemplary readers (Reader 1 and 4) for measures of drusen, loss of ellipsoid loss (EZ-loss), hyperreflective dots (HRD), hypertransmission of OCT-signal (HT), intraretinal fluid (IRF), descent of outer plexiform layer (OPL-descent), pigment epithelial detachment (PED), loss of retinal pigment epithelium (RPE-loss), subretinal fluid (SRF), and subretinal hyperreflective material (SRHM). The measurement differences (diff.) are plotted against their mean. The solid line indicates the mean difference and the dashed lines indicate the 95% limits of agreement. There were no systematic differences between the readers. Bland-Altman plots for the inter-reader agreement between each pair of all readers can be found in the Supplementary Figures S2 - S11.

More reliable than size, extent, or number of annotated features, the Dice-coefficients revealed consistent values over 0.5 (up to >0.75, Table 3) for all features. This indicated a distinct overlap of annotated regions and therefore uniform localization of the features (Figure 1).

**Table 3:**
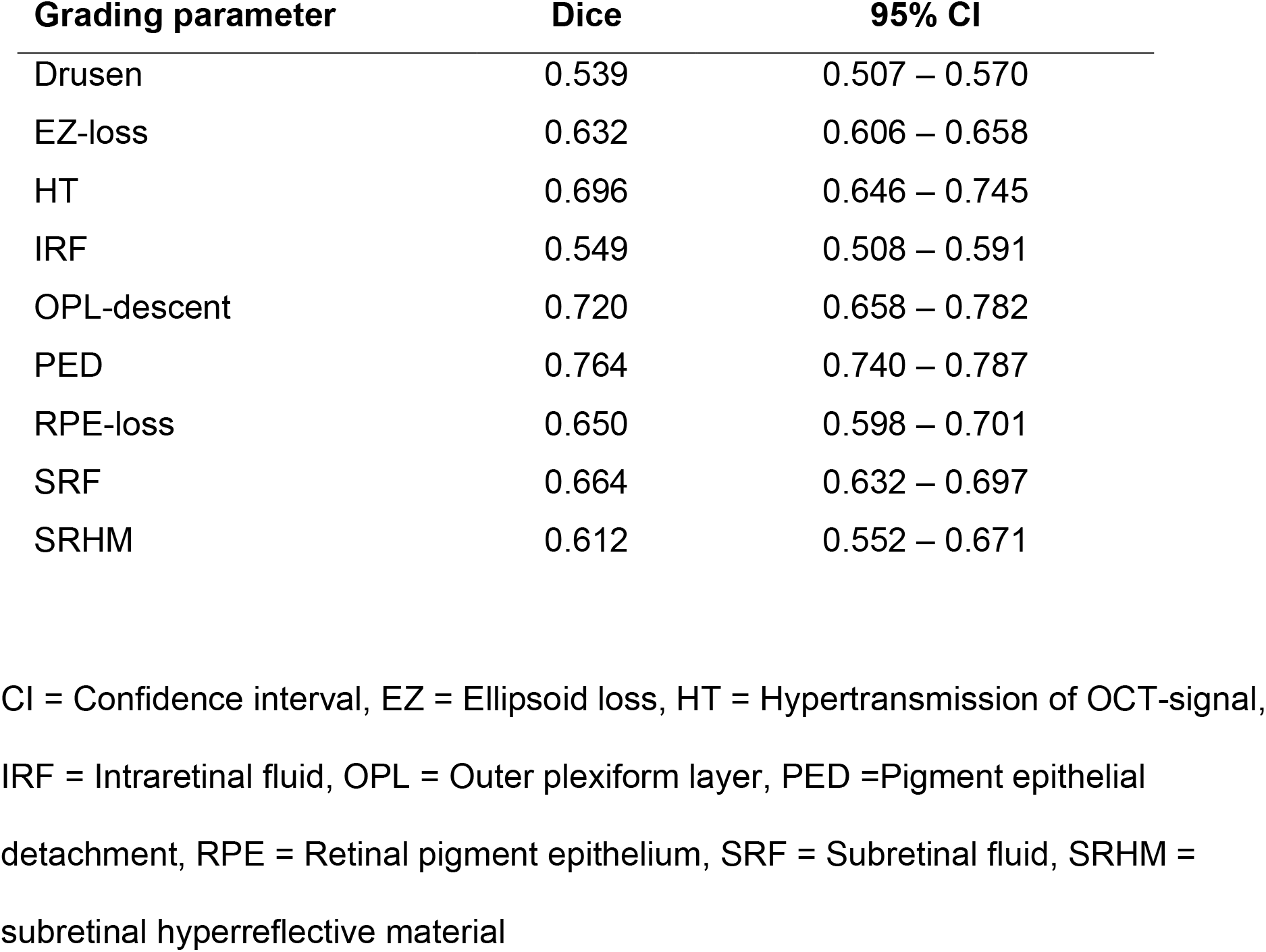
Inter-reader agreement of location of annotated features.

## DISCUSSION

In this study, we systematically investigated the reliability of grading an extensive number of structural OCT features associated with different stages of AMD in a reading center setting. The presented findings provided evidence for the dependence of inter-reader agreement on the respective annotated feature. Hence, the appropriate selection of features has the potential to provide suitable surrogate markers for disease progression and possible therapeutic effects on different disease stages in upcoming interventional trials.

Clinical surrogate markers are needed to accelerate future interventional trials. Best corrected visual acuity loss does not always constitute a useful endpoint in clinical trials for AMD due to its high interindividual variability, its psychophysical nature and phenomena such as foveal non-involvement.^33^ Nevertheless, most interventional trials for neovascular AMD currently rely on this feature. In contrast, studies for dry AMD usually use morphologic endpoints like RORA (e.g. by semiautomated delineation in fundus autofluorescence imaging)^34^ as an accepted endpoint by regulators.^8,9^ However, RORA represents the end-stage of AMD and the most effective upcoming therapeutic approach might be directed to earlier disease stages, which is difficult to extrapolate from preclinical data.^10^ Ideal surrogate markers, therefore, should (I) be readily captured, (II) reflect the current disease stage, (III) be reliable, and (IV) ideally be predictive for long-term progression based on short-term changes.^35^

As the OCT is the most abundant digital imaging device in modern ophthalmology, it has already been implemented in routine patient assessment and most clinical trial designs for retinopathies.^36^ For neovascular AMD, the analysis of IRF and SRF is used to evaluate disease activity and treatment indication besides drop of vision, presence of bleedings or leakage in angiography.^21,37^ It has been shown to be an objective and susceptible measure that might even precede functional impairment and be faster executed and/or more comfortable than invasive imaging technology like angiography or fundus photography.^21,22,38^ For dry AMD, multimodal assessment (including OCT) of drusen, pigment epithelial alterations or signs of RORA is inevitable in the differential diagnosis and analysis of disease progression.^15^ The evaluation of additional or individual OCT features could therefore be effectively carried out.

A current publication showed a consistent sequence of OCT-features in the development of RORA secondary to Maternally inherited diabetes and deafness (MIDD), indicating that these features represent different disease stages.^12^ Given that MIDD is a mitochondriopathy and mitochondrial dysfunction is considered part of the pathophysiology in AMD, results obtained in that model disease might be partly transferred to AMD. Indeed, an international consensus published by the CAM group indicated most of these features to be associated RORA development secondary to AMD.^13,14^ It also described features like EZ-loss, RPE-loss, HT, OPL-descent, HRD, and SRHM. However, the reliability of these features has not yet been comprehensively investigated by this group.

Reliability might be the most important prerequisites to define a surrogate marker for patient assessment and future interventional clinical trials. Rather low inter-reader agreement was found in the detection of the features EZ-loss and RPE-loss. Reliability of size and location of both feature annotations, however, were distinctly higher, while ICC did not reach levels of previous published data (0.75 for RPE-loss).^39^ However, the latter uses another OCT device (Spectralis HRA-OCT) that might have led to better image quality. Some of the differences between readers might be due to inaccurate delineation of lesion borders as loss and attenuation of RPE and/or EZ might merge (Figure 1). Interestingly, the average relative difference between two readers for RPE-loss was indicated with 72.4 which was significantly higher than the CV (44.8) in our study, while both measures are thought to be independent from lesion size. Concerning HRD, the variable number might derive from the size of the feature. Readers might have simply overlooked small features, leading to not more than moderate reliability (Figure 1). In this context, an automated artificial intelligence-based feature detection might have great potential to overcome this human limitation.^40,41^ Moreover, deep learning and its broader family, machine learning, is likely to be the only way to quantitate large volumes of dense OCT raster scans that are being generated in clinical trial reading centers, busy clinical practices and emerging home/remote OCT devices.^42^ However, the machine learning algorithms will be trained and the performance will be judged by the human “gold standard”,^43^ which, if unreliable, may be problematic.

More consistent results could be found for SRF and IRF. Here, our results revealed high inter-reader agreement in all three investigated parameters (detection, size, and location, Figure 1). This was in line with previously published data.^44^ Despite different datasets, the here described ICC between readers were higher than the ICC derived from inter-modality reliability between spectral domain and time domain OCT.^45,46^ Given that both features reflect neovascular activity and guide the indication for anti-VEGF treatment (besides other clinical features including hemorrhage and loss of vision), this might be of particular importance. A recent study has investigated the inter-reader agreement of PED size measures and reported an ICC of over 0.99.^47^ The slightly higher ICC value (our study, 0.972) might be traced back to the fact that the latter has only included 20 eye with definite presence of PED, and did not parallelly focus on other retinal alterations.

We noted a high reliability of the HT feature, supporting previously published.^39^ In contrast, no previous report has systematically investigated inter-reader agreement of OPL-descent. Given the high reliability (Table 1-3) and appearance in the development of RORA,^13^ OPL-descent would be worth further investigations and to explore its potential as possible surrogate marker in future clinical trials as well as for training machine learning algorithms.

Interestingly, the reliability of OCT-based features annotation for e.g. SRF, HT, and PED reached the reliability of RORA grading in fundus autofluorescence imaging in different diseases including AMD.^35,48–50^ However, OCT imaging uses less energetic infrared light that minimizes potential light toxicity and is more comfortable for the patient.^48,51^ Furthermore, OCT imaging does not rely on pupil dilation and devices are more common that fundus autofluorescence imaging devices.^36^ In this context, OCT-scans were selected in a randomized manner in our study. A previous study revealed that more eccentric scan locations might lead to less reliable results.^44^ Therefore, the pure evaluation of central scans might have led to even higher inter-reader agreement. Nevertheless, additional features of summation images like shape-descriptive parameters or dynamic flow signal could give further information,^49,50,52^ suggesting a multimodal assessment as gold standard in AMD diagnosis and study design at the current stage of imaging technology.^15^

It has been shown by the AREDS study that the number and size of drusen might predict progression of AMD.^53^ Also, the presentation of HRD,^54^ and the size of baseline RORA^55^ was reported to affect future progression rate. Concerning exudative complications, the predictive value of SRF has been controversially discussed,^56,57^ while the extent of central retinal thickening and IRF and is thought to represent the neovascular activity and therefore visual outcome.^58–60^ Therefore, it might be hypothesized that some of the additionally presented imaging features could also be predictive either for neovascular or dry AMD progression. However, the image feature description in this study was based on retrospective cross-sectional data, as it was beyond the scope of this study to evaluate the accuracy of predictive factors. However, the uniformity of the detection, size and location of most imaging features have the potential to provide the framework for further prospective studies. These prospective studies would allow to further evaluate the predictive value, which might give more insights into the pathophysiology of AMD and allow for effective study design as presented before for different parameters in AMD or other retinopathies.^38,49,50,52,61,62^ A further limitation of this study is the application of OCT imaging devices by a single manufacturer. Different OCT imaging devices might provide different scanning artefacts or image quality.^63,64^ Thereby the annotation and, hence, the reliability of single features might be different on large-scale real-world data.^65^ As there is no gold standard, it cannot be excluded that features have been missed and other data sets could provide addition conclusions. To minimize this possibility, we relied on trained retinal specialists that have identified and interpreted the features and the opportunity to add additional features was given at all timepoints during annotation (Supplementary Figure S1).

In conclusion, this study evaluated the reliability of annotations of multiple OCT features representing different disease stages in a reading-center setup. The inclusion of objective and reliable features like SRF, IRF, HT, OPL-descent or PED into future studies might enable multiple surrogate markers representing different disease stages within a single image. This might open up numerous new opportunities for evaluating disease progression and possible treatment effect in AMD, possibly leading to a more time- and cost-effective interpretation, further insights into the pathomechanisms, and improved individualized patient assessment.

## Data Availability

Data might be made available upon serious request.

## ACKNOWLEDGEMENTS

This work was supported by the German Research Foundation (grant # MU4279/2-1 to PLM), the United Kingdom’s National Institute for Health Research of Health’s Biomedical Research Centre for Ophthalmology at Moorfields Eye Hospital and UCL Institute of Ophthalmology. The views expressed are those of the authors, not necessarily those of the Department of Health. The funder had no role in had no role in the design and conduct of the study; collection, management, analysis, and interpretation of the data; preparation, review, or approval of the manuscript; and decision to submit the manuscript for publication.

## Supplementary Material

**Supplementary Figure S1:**
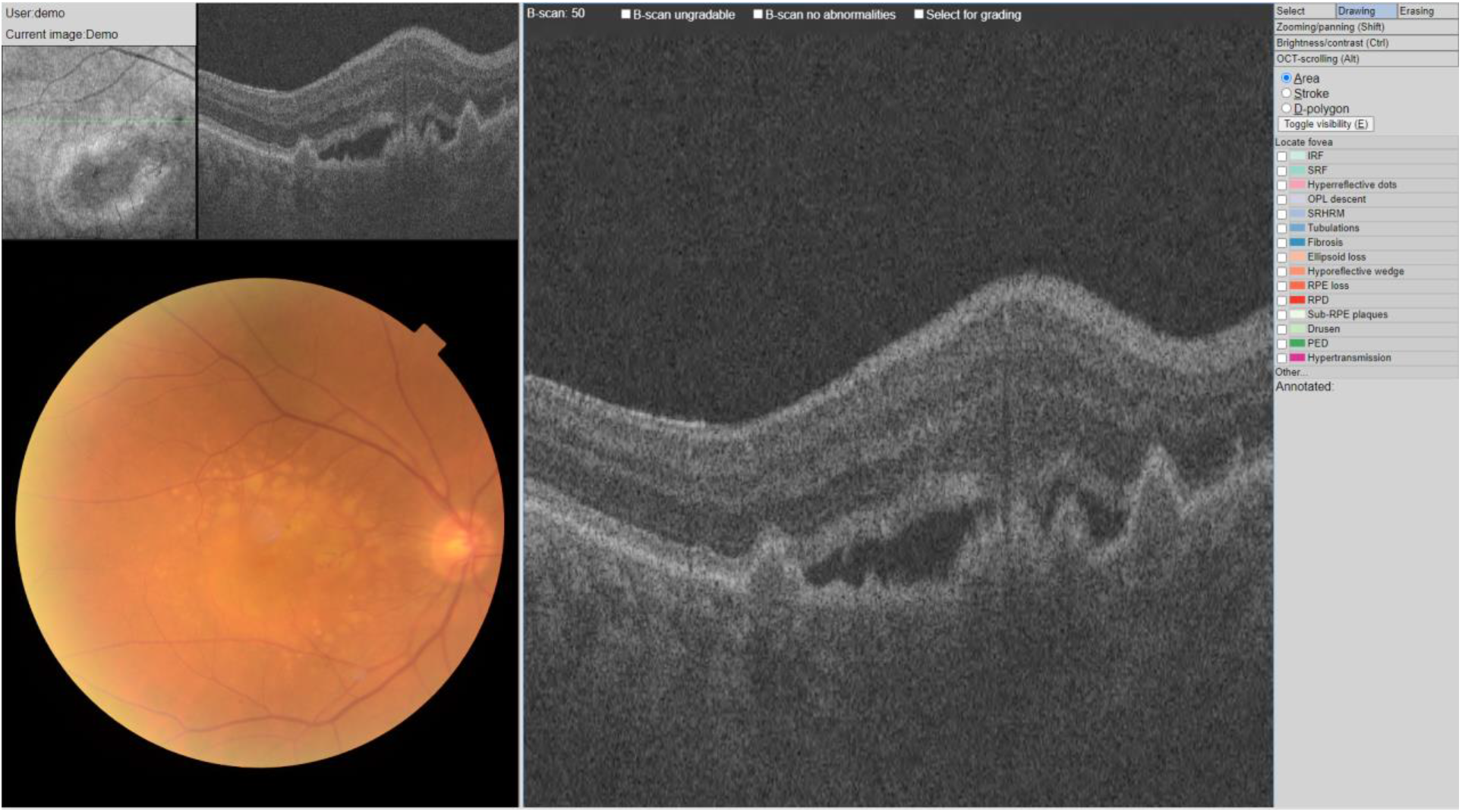
Annotation tool. The custom-build platform for annotations is demonstrated. The annotations were made in the highlighted optical coherence tomography (OCT) B-scan (middle) by choosing the respective label (right). Additional labels not covered by the default setup could be added under “Other…”. According to the label, the features were either annotated as area, stroke (i.e., line or point), or D-polygon. Wrong annotations could be deleted by selecting “Erasing” in the top-right corner. Ungradable B-scans or those without abnormalities could be labeled using the boxes at the top. On the top left the infrared reflectance image showed the location of the respective B-scan as green line next to a small preview of the B-scan. During the annotations, the reader could scroll through the other scans of the eye for context. The according color fundus photography was visualized at the bottom left.

**Supplementary Figure S2:**
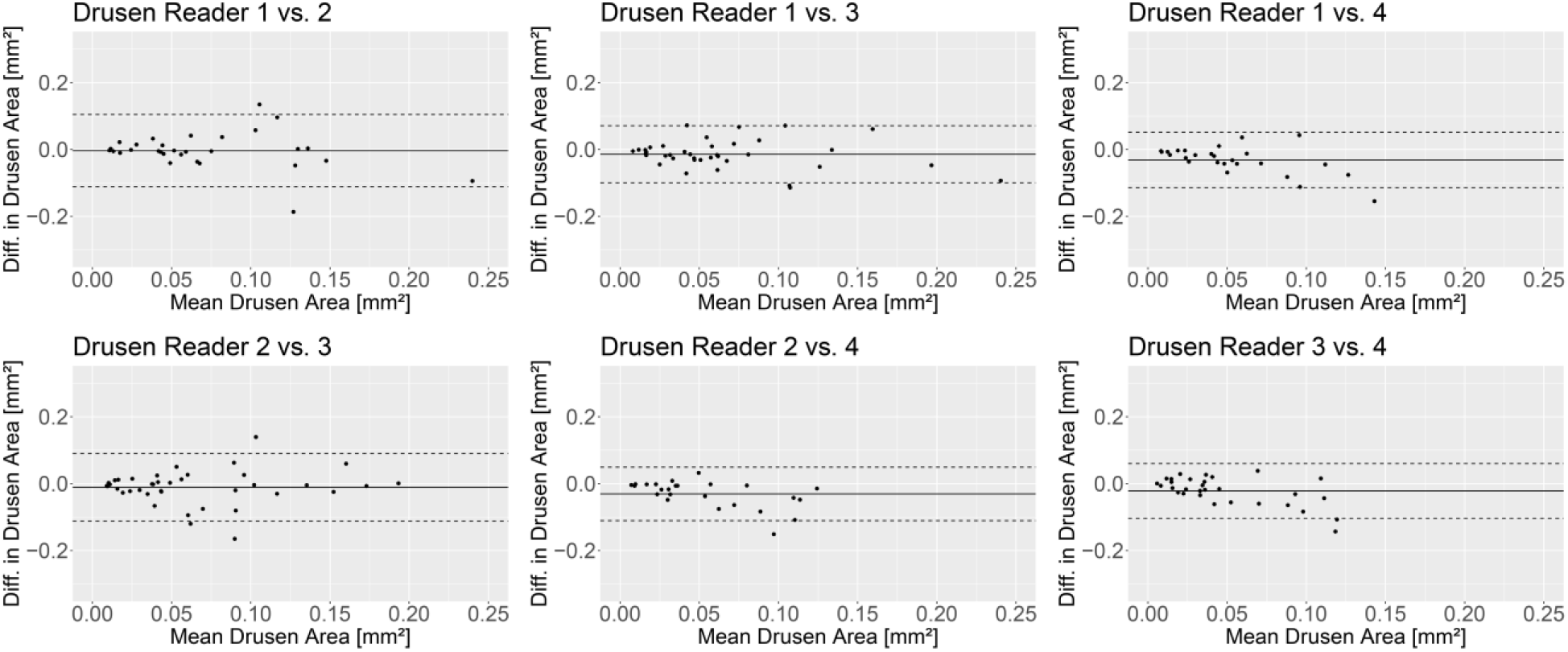
Inter-reader agreement for the measures of the area of drusen. The Bland-Altman plots demonstrate the measurement differences (diff.) of two readers plotted against their mean. The solid line indicates the mean difference and the dashed lines indicate the 95% limits of agreement. There were no systematic differences between the readers.

**Supplementary Figure S3:**
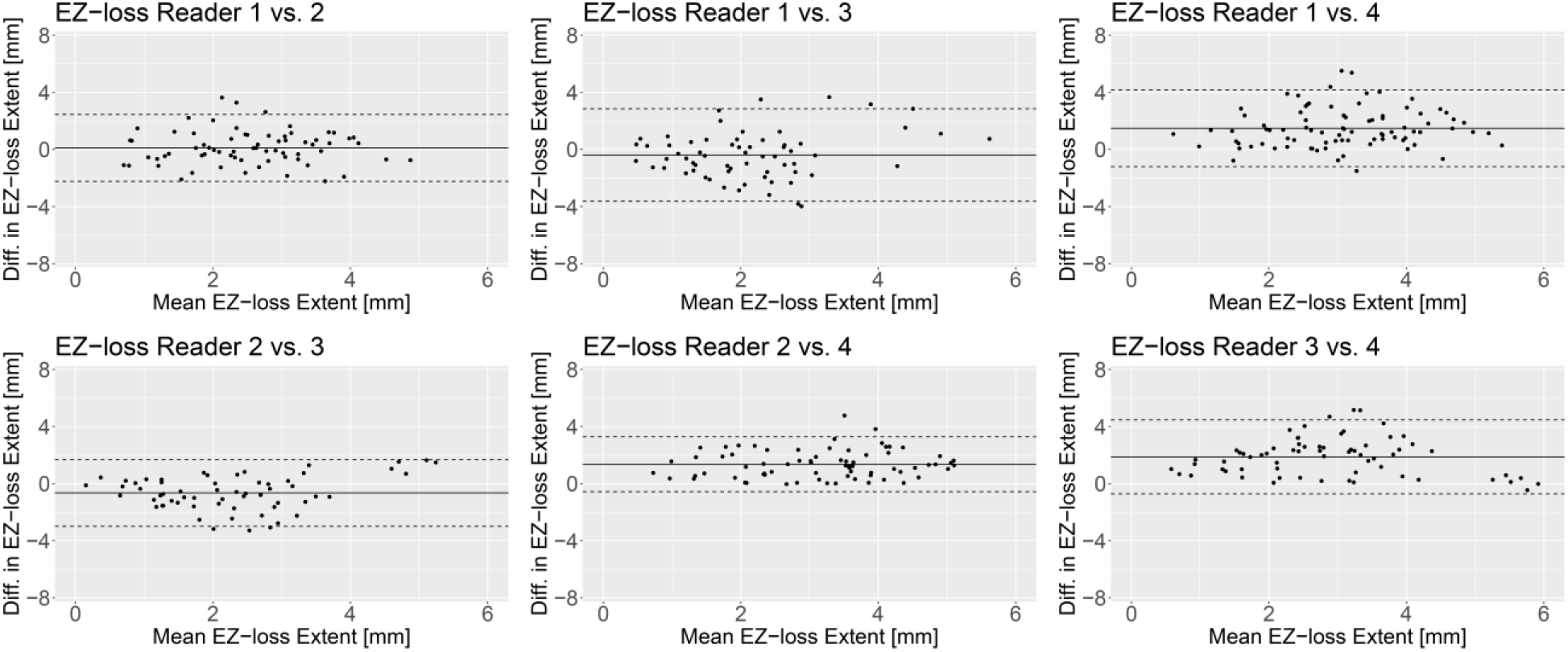
Inter-reader agreement for the measures of the extent of ellipsoid zone (EZ) loss. The Bland-Altman plots demonstrate the measurement differences (diff.) of two readers plotted against their mean. The solid line indicates the mean difference and the dashed lines indicate the 95% limits of agreement. There were no systematic differences between the readers.

**Supplementary Figure S4:**
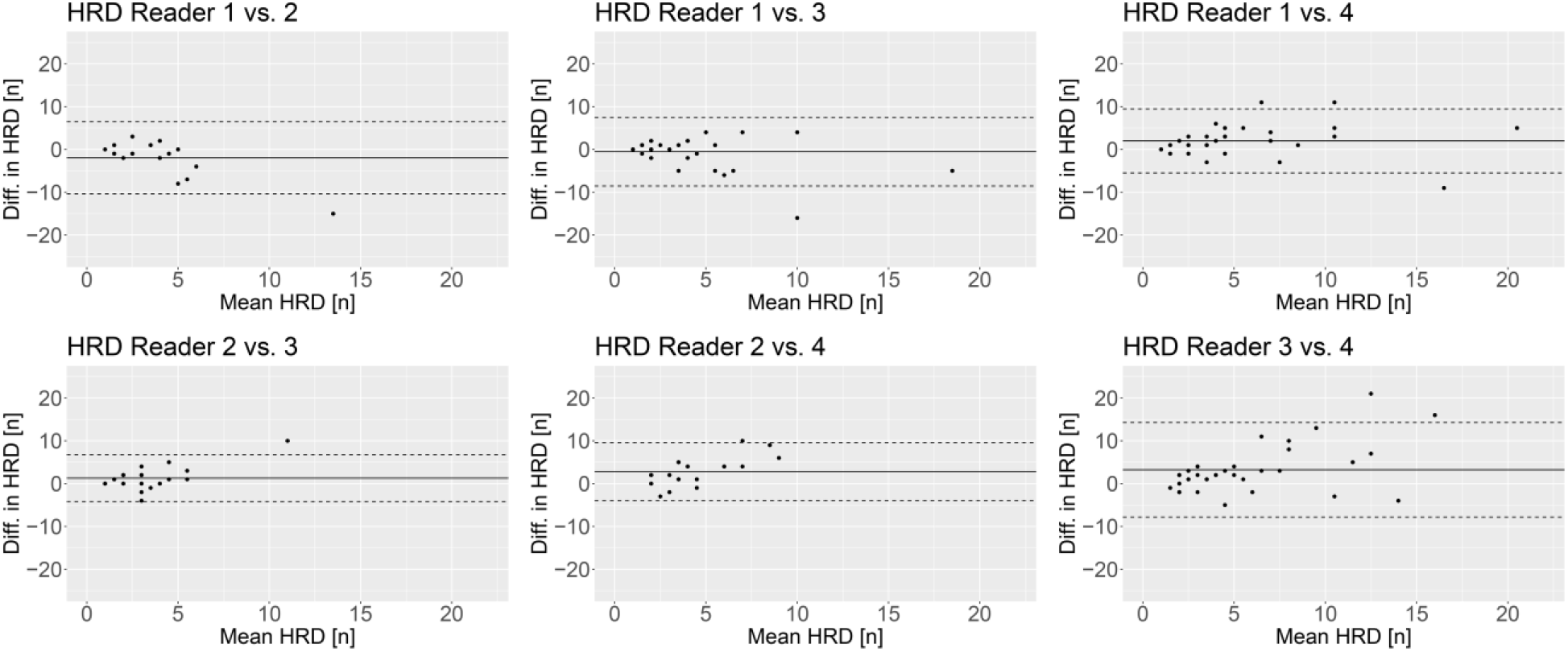
Inter-reader agreement for the measures of hyperreflective dots (HRD) The Bland-Altman plots demonstrate the measurement differences (diff.) of two readers plotted against their mean. The solid line indicates the mean difference and the dashed lines indicate the 95% limits of agreement. There were no systematic differences between the readers.

**Supplementary Figure S5:**
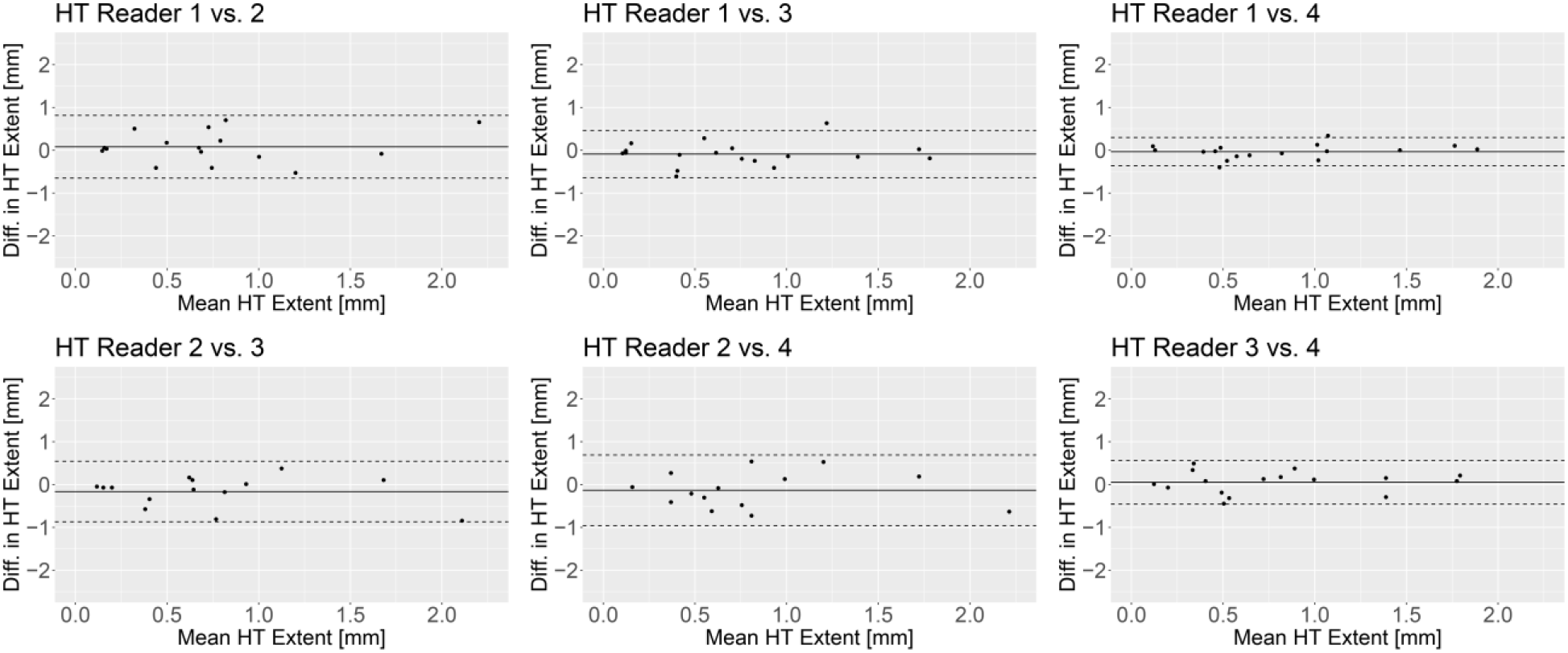
Inter-reader agreement for the measures of the extent of OCT signal hypertransmission (HT) The Bland-Altman plots demonstrate the measurement differences (diff.) of two readers plotted against their mean. The solid line indicates the mean difference and the dashed lines indicate the 95% limits of agreement. There were no systematic differences between the readers.

**Supplementary Figure S6:**
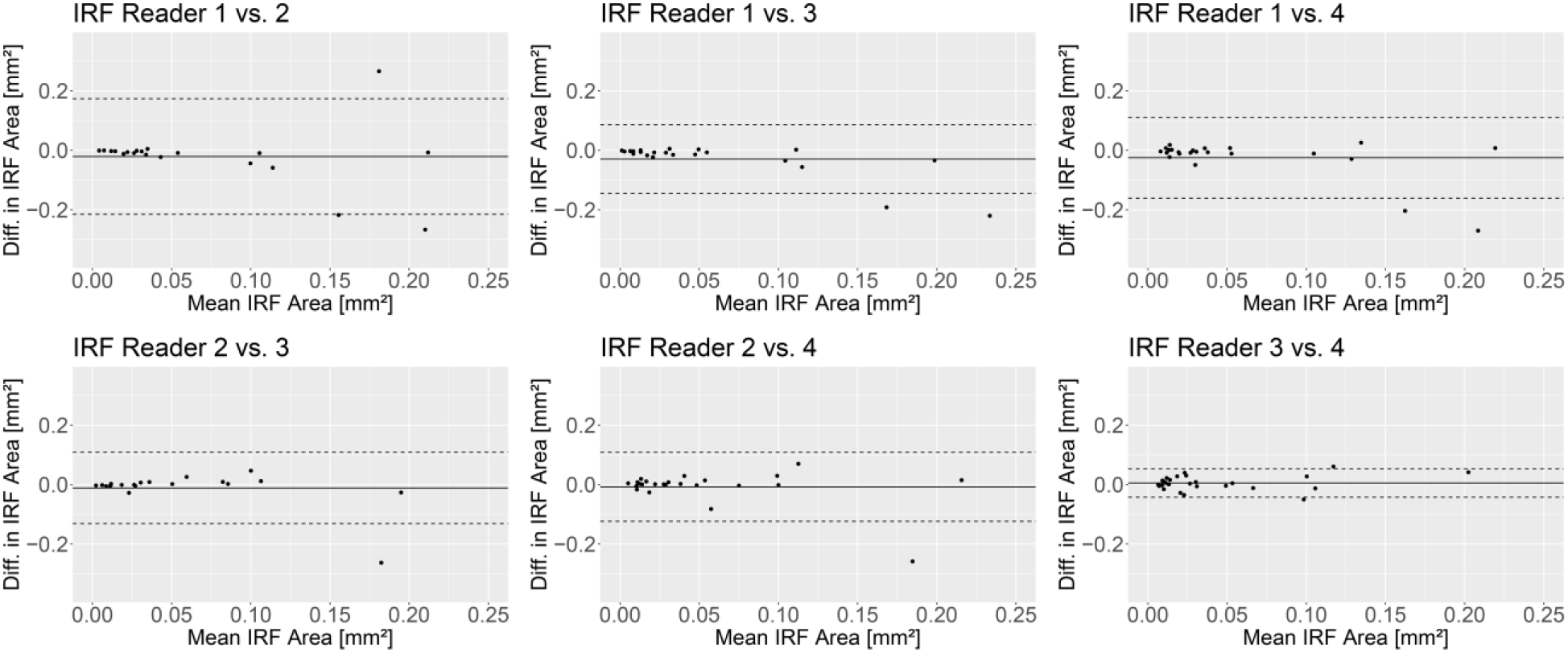
Inter-reader agreement for the measures of the area of intraretinal fluid (IRF) The Bland-Altman plots demonstrate the measurement differences (diff.) of two readers plotted against their mean. The solid line indicates the mean difference and the dashed lines indicate the 95% limits of agreement. There were no systematic differences between the readers.

**Supplementary Figure S7:**
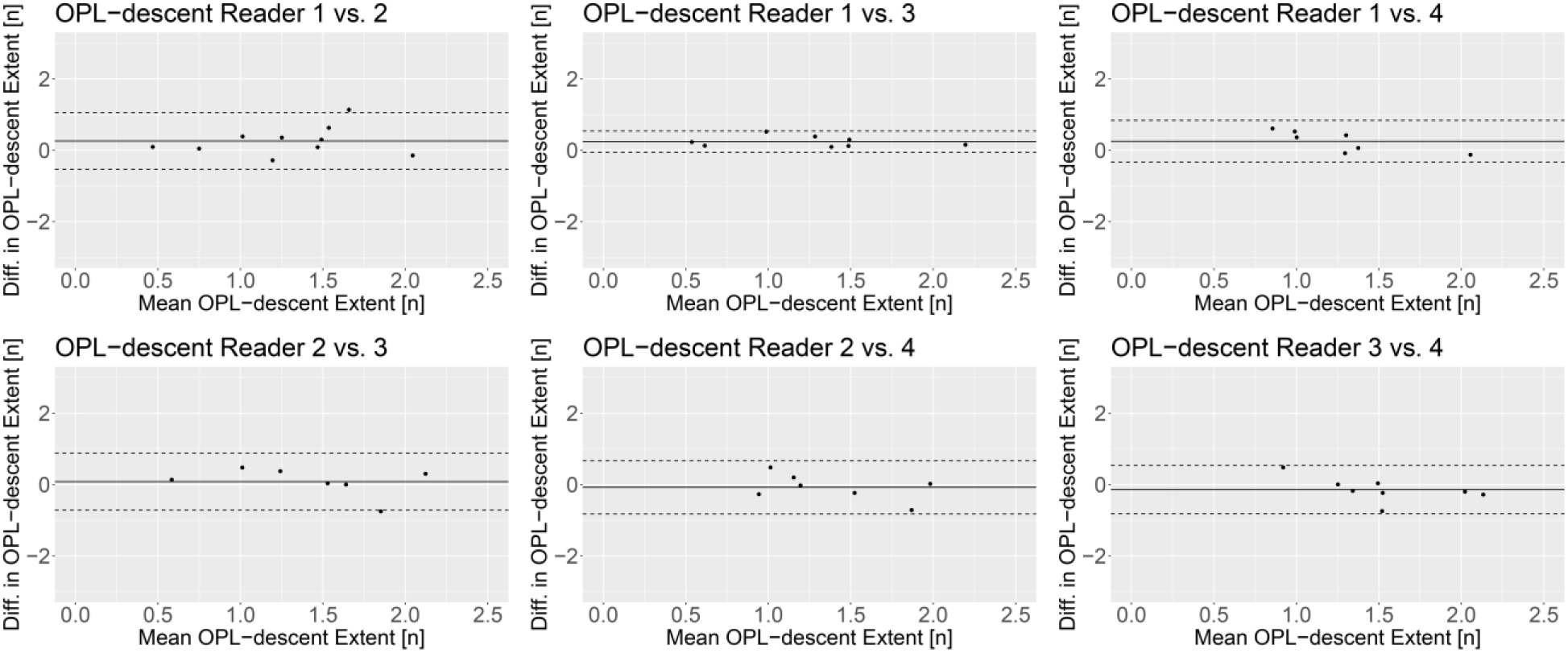
Inter-reader agreement for the measures of the extent of outer plexiform layer (OPL) descent. The Bland-Altman plots demonstrate the measurement differences (diff.) of two readers plotted against their mean. The solid line indicates the mean difference and the dashed lines indicate the 95% limits of agreement. There were no systematic differences between the readers.

**Supplementary Figure S8:**
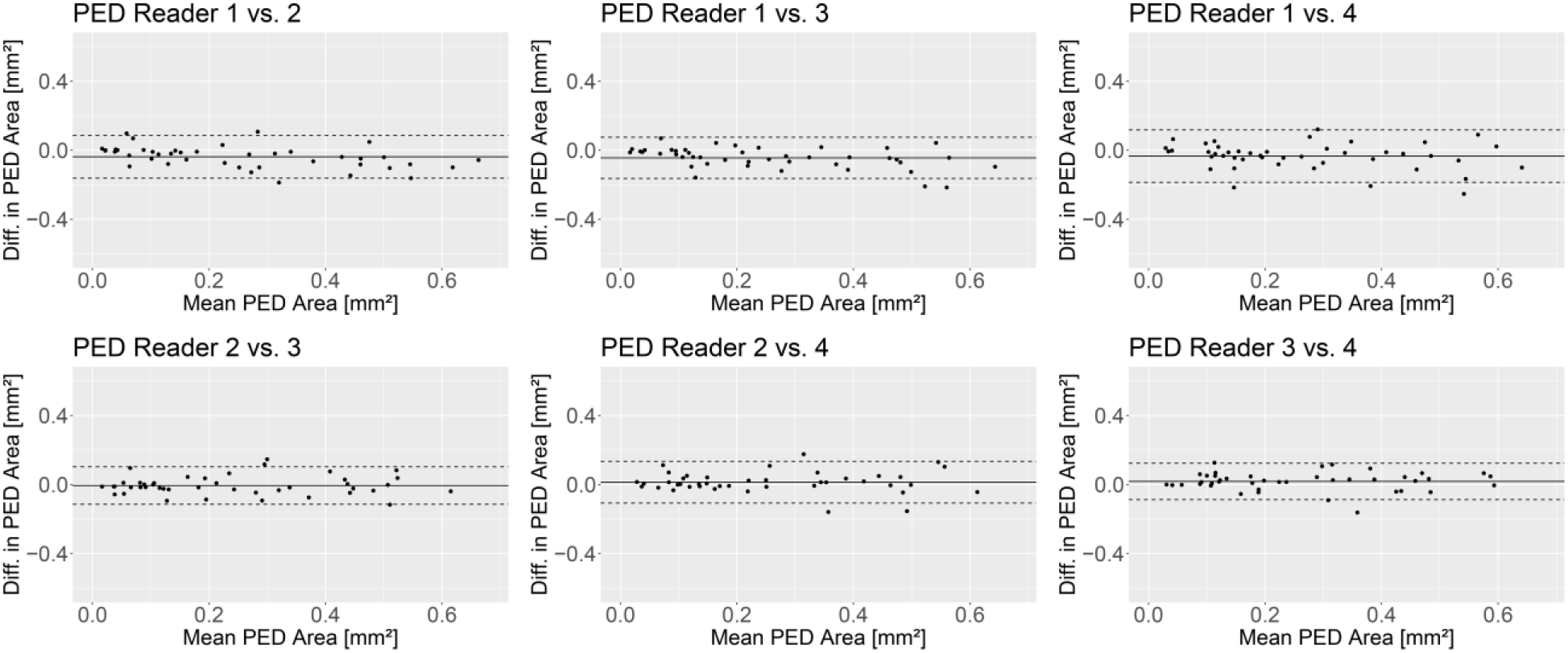
Inter-reader agreement for the measures of the area of pigment epithelial detachment (PED) The Bland-Altman plots demonstrate the measurement differences (diff.) of two readers plotted against their mean. The solid line indicates the mean difference and the dashed lines indicate the 95% limits of agreement. There were no systematic differences between the readers.

**Supplementary Figure S9:**
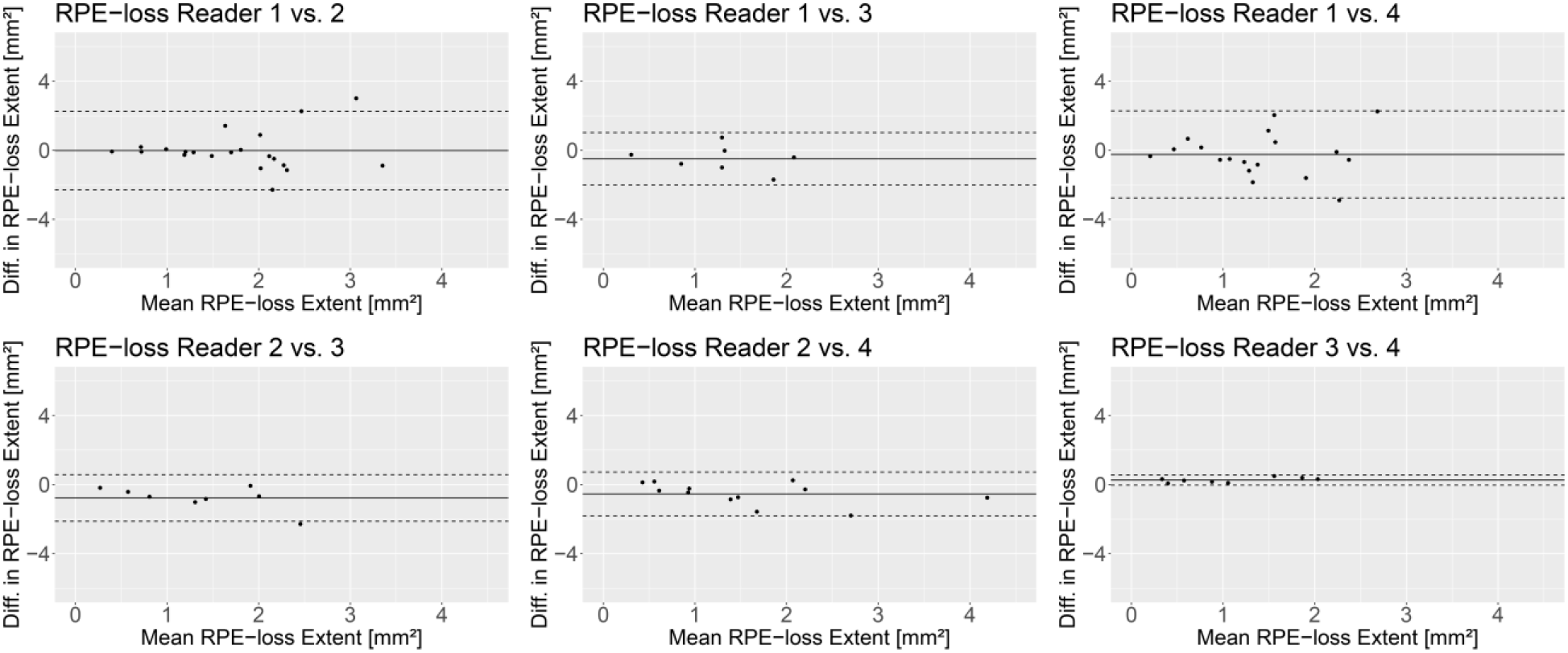
Inter-reader agreement for the measures of the extent of retinal pigment epithelium (RPE) loss. The Bland-Altman plots demonstrate the measurement differences (diff.) of two readers plotted against their mean. The solid line indicates the mean difference and the dashed lines indicate the 95% limits of agreement. There were no systematic differences between the readers.

**Supplementary Figure S10:**
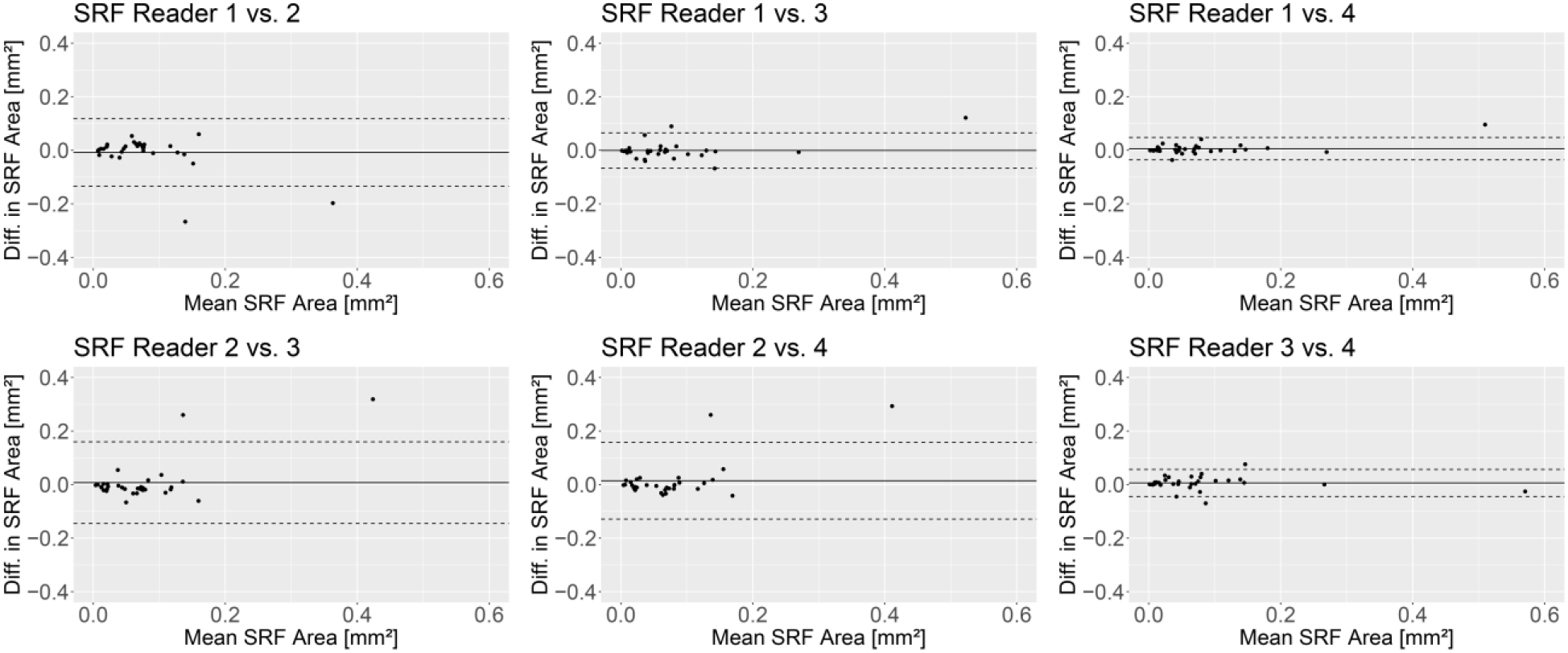
Inter-reader agreement for the measures of the area of subretinal fluid (SRF) The Bland-Altman plots demonstrate the measurement differences (diff.) of two readers plotted against their mean. The solid line indicates the mean difference and the dashed lines indicate the 95% limits of agreement. There were no systematic differences between the readers.

**Supplementary Figure S11:**
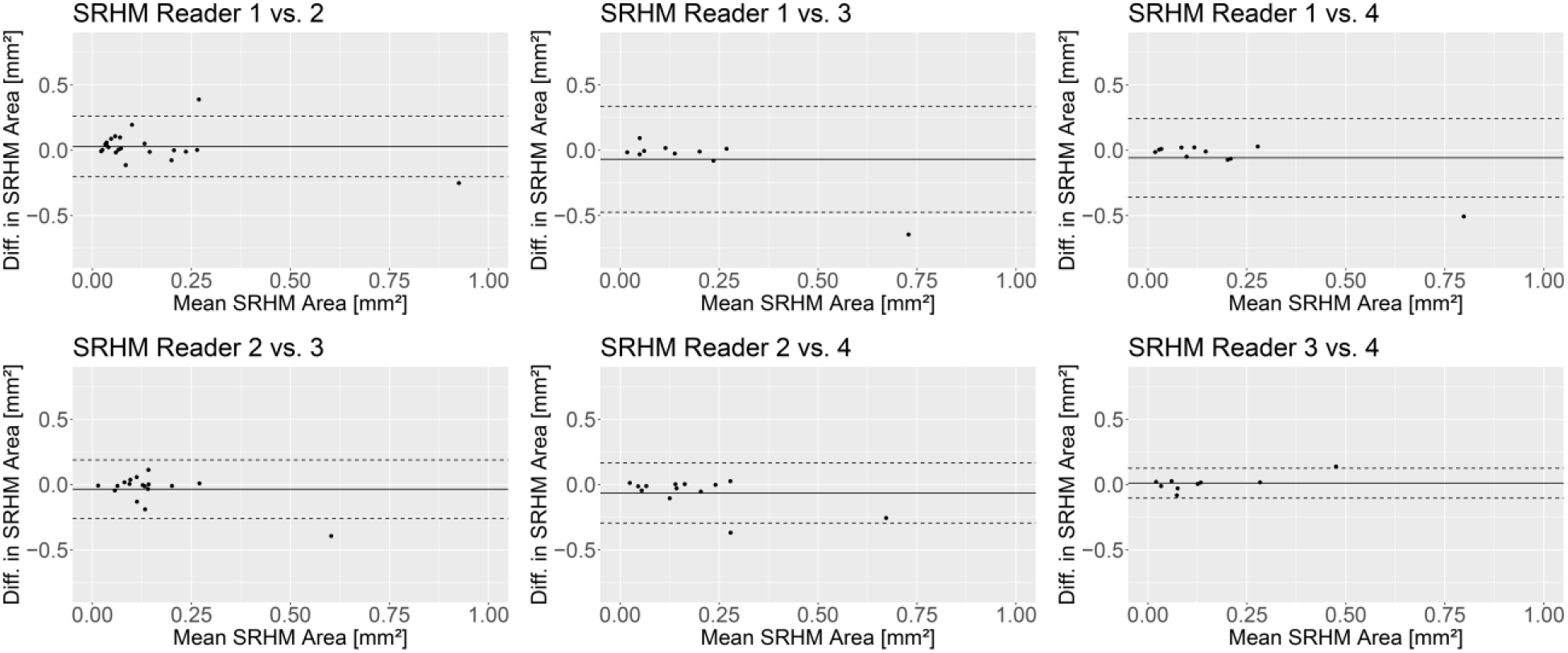
Inter-reader agreement for the measures of the area of subretinal hyperreflective material (SRHM) The Bland-Altman plots demonstrate the measurement differences (diff.) of two readers plotted against their mean. The solid line indicates the mean difference and the dashed lines indicate the 95% limits of agreement. There were no systematic differences between the readers.

